# Polygenic Risk Scores for Cardiovascular Disease Predict Risk Factor Control and Residual Cardiovascular Risk in Stroke Survivors

**DOI:** 10.64898/2026.08.18.26360673

**Authors:** Nicola Luigi Bragazzi, Lanyue Zhang, Murad Omarov, Luka Živković, Marios K Georgakis

## Abstract

**Background:** Stroke remains a leading cause of mortality and long-term disability worldwide, with high residual vascular risk among survivors despite optimal secondary prevention. The contribution of inherited polygenic risk to this residual vulnerability remains unclear.

**Methods:** We analyzed 2,701 stroke survivors (mean age 59.8±7.1 years, 61.7% male) from the UK Biobank. Stroke- and coronary artery disease (CAD)-polygenic risk scores (metaGRS), comprising approximately 3.2 million and 1.7 million genetic variants, respectively, were derived from large-scale genome-wide association studies using penalized regression. The primary outcome was major adverse cardiovascular events (MACE), while secondary outcomes included recurrent stroke and vascular risk factor control. metaGRS associations with incident MACE and recurrent stroke were tested using Cox models, whereas associations with baseline risk-factor control were assessed using logistic regression. Mediation analyses quantified indirect effects of metaGRS to MACE *via* HbA1c, LDL cholesterol, and blood pressure.

**Results:** Over 12 years, 731 MACE events (27.1%) and 351 recurrent stroke events (13.0%) occurred. CAD-metaGRS was independently associated with future MACE (age- and sex-adjusted HR per SD increment 1.15, 95%CI 1.07–1.24; p<0.001), whereas higher stroke- and CAD metaGRS were both associated with poorer glycemic control. A higher CAD-metaGRS was also associated with poorer lipid control. Mediation analyses identified glycemic regulation as a significant pathway linking polygenic risk to recurrent vascular events.

**Conclusions:** Polygenic risk scores for cardiovascular disease are associated with recurrent vascular events and vascular risk factor control among stroke survivors, pointing to potentially actionable insights in secondary prevention that merit further investigation.

**Graphical abstract.:** 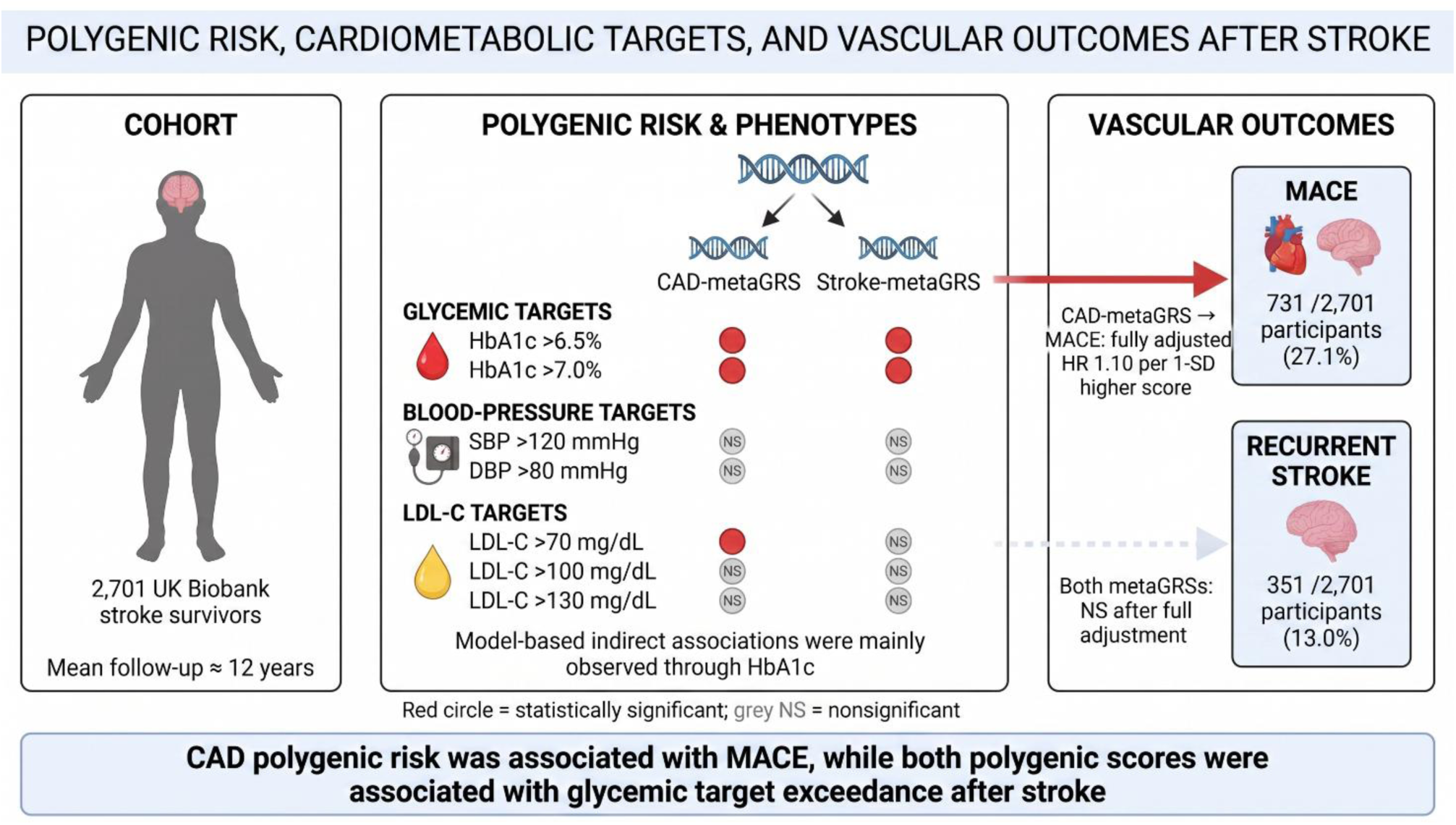

## Introduction

Stroke is one of the leading causes of mortality and long-term disability worldwide, accounting for over 6.6 million deaths annually and ranking as the second leading cause of death and third leading cause of disability-adjusted life years globally.^1,2^ Advances in acute management have markedly improved survival, leading to a steadily growing population of stroke survivors.^3,4^ However, this population faces substantial residual vascular risk despite adherence to evidence-based pharmacologic and lifestyle management. Contemporary cohort data show that among patients discharged after ischemic stroke (IS) or transient ischemic attack, the 1-year cumulative incidence of major adverse cardiovascular events (MACE), including recurrent IS, myocardial infarction (MI), and vascular death, remains approximately 12–13%, rising to over 35% within five years of follow-up.^5–7^ Even under optimal treatment, the 5-year risk of recurrent IS alone ranges from 10% to 16%, underscoring a persistent biological vulnerability not fully explained by conventional risk factors.^8,9^

Understanding and quantifying this residual risk represents a critical frontier in secondary stroke prevention. A key component of this residual vulnerability likely resides in inherited predisposition. Genome-wide association studies (GWAS) have elucidated the polygenic architecture of both stroke and coronary artery disease (CAD), identifying thousands of common variants that collectively contribute to vascular susceptibility.^10–12^ Stroke survivors are at risk of both recurrent cerebrovascular and coronary events; therefore, genetic liability to CAD may be relevant after an index stroke.^5,9^ Polygenic or genetic risk scores (GRS), which aggregate these effects across the genome, have emerged as powerful instruments to summarize cumulative genetic liability.^13^ A metaGRS can combine multiple phenotype-specific scores and millions of variants weighted by their GWAS effect sizes, thereby refining individualized risk estimation.^14^ However, despite extensive validation in primary prevention cohorts, their relevance in secondary prevention, particularly among stroke survivors, remains underexplored.^13,15^ This population is pathophysiologically distinct, characterized by high comorbidity burden, pharmacologic exposure, and disease heterogeneity. Whether genetic risk retains prognostic and actionable value in this context is an open and clinically consequential question.

In the present study, we leveraged data from the UK Biobank to address this knowledge gap. We specifically evaluated two polygenic scores, capturing the genetic underpinnings of cerebrovascular and coronary atherosclerotic risk. First, we examined whether the two GRSs independently predict recurrent vascular events (MACE and stroke) among stroke survivors. Second, we assessed whether the GRSs are associated with attainment of guideline-recommended targets. We hypothesized that polygenic risk for stroke and CAD retains prognostic value for recurrent vascular events in stroke survivors, and it may provide actionable insights to guide more intensive monitoring and secondary prevention strategies.

## Material and methods

### Study Design and Population

This study utilized data from the UK Biobank,^16^ a large prospective cohort comprising over 500,000 participants aged 40–69 years recruited between 2006 and 2010. Baseline data included extensive phenotyping, biochemical assays, and genome-wide genotyping. Follow-up information was derived from electronic health records, hospital episode statistics, and mortality registries.

For the present analysis, we identified participants with a history of stroke prior to baseline assessment, including both IS and hemorrhagic subtypes. Individuals with missing genetic information, incomplete follow-up, or withdrawn consent were excluded. The resulting cohort constituted a population of stroke survivors under longitudinal observation for incident vascular events. The exact source fields and diagnostic codes used for the cohort and outcomes are provided in **Supplemental Table 1**.

### Ethics and Data Access

The UK Biobank received approval from the National Health Service North West Multi-Centre Research Ethics Committee, and all participants provided written informed consent. Data for this work were accessed under the UK Biobank application number 151281.

### Outcomes

The primary endpoint was the occurrence of MACE, defined as a composite of nonfatal MI, nonfatal stroke, or cardiovascular death. Secondary endpoints included: i) stroke events, comprising recurrent IS or hemorrhagic stroke, and ii) the control of vascular risk factors. Clinical outcomes were ascertained using International Classification of Diseases, Tenth Revision (ICD-10) codes from linked hospital admission and death registry records (**Supplemental Table 1**).

### Genotyping and Genetic Risk Score

Genotyping in the UK Biobank was performed using the UK BiLEVE/UK Biobank Axiom arrays, followed by centralized imputation to the Haplotype Reference Consortium and UK10K reference panels. Genetic quality control excluded variants with minor allele frequency (MAF)<1%, Hardy– Weinberg equilibrium p<10^-6^, or imputation INFO<0.4.

Two major polygenic constructs were investigated: i) a stroke- and ii) a CAD-metaGRS. The former was derived from the 2018 multi-ancestry stroke GWAS (n≈520,000 individuals) incorporating IS subtypes.^14^ Specifically, the stroke metaGRS integrates 19 component genomic risk scores capturing stroke and its subtypes (including IS), CAD, and genetically correlated vascular risk factors, such as systolic (SBP) and diastolic (DBP) blood pressure, lipid traits (total, low-density lipoprotein (LDL)-, and high-density lipoprotein (HDL)-cholesterol and triglycerides, or TGs), atrial fibrillation, type 2 diabetes, body mass index (BMI), smoking, and height. The final score comprises approximately 3.2 million genetic variants. Variant weights were estimated using elastic-net penalized regression in a small UK Biobank derivation cohort and subsequently validated in an independent cohort of White British participants free of prevalent stroke at baseline. The derivation set included 11,995 UK Biobank participants; because participant identifiers were unavailable, a small degree of overlap with the present study cannot be excluded. Variant weights were estimated using penalized regression to optimize predictive performance and control for multicollinearity across correlated *loci*. The CAD-metaGRS was developed using a meta-analytic framework that combined three complementary scores capturing partially independent sources of CAD-related genetic information: a 46,000-variant score derived from the cardiometabolic Metabochip array; a 202-variant score comprising variants associated with CAD at a false discovery rate <0.05 in CARDIoGRAMplusC4D; and a genome-wide polygenic score derived from the same GWAS. The component scores were standardized and combined using log-hazard-ratio weights estimated in a UK Biobank training sample of 3,000 participants (1,000 CAD cases and 2,000 controls) while accounting for correlations among the component scores. The resulting metaGRS comprised approximately 1.7 million variants. It was subsequently evaluated in an independent, held-out sample of 482,629 UK Biobank participants, including 22,242 CAD cases and 460,387 noncases.^17^ For each participant, standardized genetic scores were computed as the sum of risk allele dosages weighted by their corresponding effect sizes using PLINK 2 software, followed by z-transformation within the sample.

Distributions of the polygenic constructs and their correlations with the other variables are shown in **Supplemental Figures 1** and **2**.

### Covariates

Models were adjusted for age, sex, anthropometric variables (BMI, waist, and hip circumference), baseline hypertension, diabetes mellitus, smoking status, SBP, DBP, glycated hemoglobin (HbA1c), and lipid profile, including HDL-C, LDL-C, and TGs, as well as estimated glomerular filtration rate based on combined creatinine and cystatin C (eGFRcr-cys). Covariates were obtained from the UK Biobank baseline assessment and included self-reported medical history and lifestyle factors, standardized physical measurements, and centrally performed biochemical assays, as appropriate.

### Statistical Analysis

The analytic workflow proceeded in three stages. First, we established a clinical reference model predicting MACE, as the primary outcome, and recurrent stroke, as a secondary outcome, among stroke survivors using Cox proportional hazards regression. Model discrimination was quantified by Harrell’s C-index, with likelihood ratio test (LRT) statistics computed to evaluate relative model fit. Second, we evaluated the independent prognostic value of the stroke- and CAD-metaGRSs by fitting age- and sex-adjusted as well as fully adjusted Cox models (which included all prespecified demographic, anthropometric, biochemical, and clinical covariates), subsequently assessing their incremental predictive value beyond the clinical reference model through comparisons of discrimination and model fit. Third, associations of each metaGRS with baseline failure to achieve glycemic, blood-pressure, and lipid targets were assessed using univariable and age- and sex-adjusted logistic regression models. Results were expressed as odds ratios (ORs) with 95% confidence intervals (CIs). To elucidate dose–response relationships, participants were stratified by percentile distributions of stroke- and CAD-metaGRS. We next assessed whether GRS effects on MACE and recurrent stroke events were mediated through modifiable cardiometabolic risk factors, specifically glycemic control (HbA1c), blood pressure (SBP and DBP), and LDL-C. Vascular risk-factor thresholds were selected to reflect both historically contemporaneous secondary prevention guidance during UK Biobank recruitment and follow-up and clinically meaningful levels of control (HbA1c >6.5% and >7.0%; SBP >120 mmHg; DBP >80 mmHg; LDL-C >70, >100, and >130 mg/dL).^18,19^

Model-based, horizon-specific interventional survival mediation analyses were performed to quantify the direct and indirect genetic effects on vascular outcomes. All mediator models included standardized age and sex. Time to MACE or recurrent stroke was modelled using right-censored Weibull proportional-hazards regression including the relevant metaGRS, mediator, metaGRS-by-mediator interaction, standardized age, and sex. Model-based marginal risks at 5, 10, and 15 years were obtained by parametric g-computation and standardized over the empirical age and sex distribution of the cohort. Binary-mediator distributions were integrated exactly, whereas continuous-mediator distributions were integrated using 20-point Gauss–Hermite quadrature. At each horizon, the total association was decomposed into interventional direct and indirect components, expressed as absolute risk differences in percentage points. The 10-year horizon was designated as primary, with the 5- and 15-year estimates treated as secondary; the 15-year estimates were considered extrapolative because fewer than 7% of participants remained under observation at that horizon. Uncertainty was quantified using 5,000 participant-level nonparametric bootstrap resamples per mediator–pathway model, with both the mediator and outcome models refitted in every resample. The same bootstrap fit generated estimates at all three horizons. We calculated 95% percentile bootstrap CIs and two-sided empirical bootstrap p-values using a +1 correction. Benjamini–Hochberg false-discovery-rate correction was applied to the indirect-effect p-values separately within each pathway and time horizon across the 11 mediator specifications.

All analyses were conducted in R version 4.5.3 (R Foundation for Statistical Computing, Vienna, Austria). Cox proportional hazards models were fitted using the *survival* package, whereas logistic regression models were fitted using the *stats* package. Concordance indices were computed using the *Hmisc* package, LRT via *rms*, paired differences in Harrell C-indices via infinitesimal-jackknife covariance estimates from the *concordance* function (*survival* package), two-sided Wald 95%CIs and p-values via the extended-minus-reference contrast, and model-based, horizon-specific interventional survival mediation analyses through the *lm.fit* and *optim* functions from the *stats* package.

## Results

### Study Population and Baseline Characteristics

The study included 2,701 stroke survivors (mean age 59.8±7.1 years; 61.7% male). Participants exhibited a substantial burden of cardiometabolic risk factors, with high prevalence of hypertension (66.3%), diabetes mellitus (13.3%), previous smoking (44.8%), antihypertensive medication use (60.0%), and statin therapy (63.7%). Mean BMI was 28.7±5.0 kg/m², mean SBP/DBP were 141.6±20.0/82.1±11.3 mmHg, and mean HbA1c was 39.1±13.1 mmol/mol (5.7±1.2%). Additional anthropometric, biochemical, and renal characteristics are summarized in **Table 1**.

**Table 1.** Baseline clinical, anthropometric, and biochemical characteristics of stroke survivors from the UK Biobank.

| Variable | Overall<br>N = 2,701 |
| --- | --- |
| Sex |  |
| Female (N, %) | 1,035 (38%) |
| Male (N, %) | 1,666 (62%) |
| Age (years) | 59.8 (7.1) |
| Waist circumference (cm) | 95.9 (13.6) |
| Hip circumference (cm) | 104.7 (9.5) |
| BMI (kg/m<sup>2</sup>) | 28.7 (5.0) |
| Systolic blood pressure (mmHg) | 141.6 (20.0) |
| Diastolic blood pressure (mmHg) | 82.1 (11.3) |
| LDL-C (mmol/L) | 3.0 (0.9) |
| (mg/dL) | 114.6 (34.5) |
| HDL-C (mmol/L) | 1.3 (0.4) |
| (mg/dL) | 51.6 (14.1) |
| Triglycerides (mmol/L) | 1.8 (1.1) |
| (mg/dL) | 162.8 (94.3) |
| Total cholesterol (mmol/L) | 4.9 (1.2) |
| (mg/dL) | 189.7 (46.2) |
| eGFR<sub>cr-cys</sub> (mL/min/1.73 m<sup>2</sup>) | 86.9 (17.6) |
| Cystatin C (mg/L) | 1.0 (0.3) |
| Creatinine (μmol/L) | 78.8 (25.3) |
| (mg/dL) | 0.9 (0.3) |
| HbA1c (mmol/mol) | 39.1 (13.1) |
| (%) | 5.7 (1.2) |
| Previous smoke | 1,209 (45%) |
| Current smoke | 409 (15%) |
| Anti-hypertensive drug usage | 1,621 (60%) |
| Statin usage | 1,719 (64%) |
| Diabetes | 360 (13%) |
| Hypertension | 1,791 (66%) |

Values are mean (SD) or n (%).

Over a mean follow-up of 4375±1355 days (12.0±3.7 years) for MACE and 4697±1075 days (12.9±2.9 years) for recurrent stroke, a total of 731 MACE events (27.1%) and 351 recurrent stroke events (13.0%) were observed.

When participants were stratified by MACE status (**Supplemental Table 2**), those who developed MACE were older and more frequently male and had greater adiposity, higher SBP, lower DBP, poorer renal function and glycemic control, and lower LDL-C, HDL-C, and total cholesterol. They also had higher prevalences of current smoking, hypertension, diabetes, antihypertensive treatment, and statin use. Participants with recurrent stroke were older and had greater adiposity, higher SBP, poorer renal function and glycemic control, and lower LDL-C, HDL-C, and total cholesterol. Antihypertensive treatment, statin use, diabetes, and hypertension were also more frequent among participants with recurrent stroke.

### Predictors of MACE and Recurrent Stroke: Cox Proportional Hazards Analysis

In the multivariable Cox model (**Supplemental Figure 3**), older age, current smoking, diabetes mellitus, impaired renal function, and higher systolic blood pressure independently predicted MACE. Specifically, current smoking (hazard-ratio, HR=1.71, 95%CI 1.38–2.12; p<0.001) and diabetes mellitus (HR=1.80, 95%CI 1.49–2.19; p<0.001) were the strongest risk factors, while each one-year increase in age was associated with a 5% higher risk (HR=1.05, 95%CI 1.03–1.06; p<0.001). Lower eGFRcr-cys (HR per 1-mL/min/1.73 m² increase=0.99, 95%CI 0.98–0.99; p<0.001), greater waist circumference (HR per 1-cm increase=1.01, 95%CI 1.00–1.03; p=0.028), and higher SBP (HR per 1-mmHg increase=1.01, 95%CI 1.00–1.01; p=0.018) were also independently associated with MACE. Statin therapy showed a modest positive association (HR=1.26, 95%CI 1.04–1.52; p=0.019), likely reflecting indication bias. No independent associations were observed for sex, hypertension, previous smoking, BMI, lipid fractions, HbA1c, DBP, or antihypertensive treatment. For recurrent stroke (Supplemental Figure 4), age and diabetes were the only independent predictors. Diabetes was associated with 77% higher risk of recurrent stroke (HR=1.77, 95%CI 1.34–2.33; p<0.001), while each additional year of age with 4% higher risk (HR=1.04, 95%CI 1.02–1.06; p<0.001). No significant associations were observed for smoking status, sex, anthropometric measures, renal function, lipid profile, HbA1c, blood pressure measurements, or medication use.

In age- and sex-adjusted Cox models, higher CAD-metaGRS (HR per SD increment=1.15; 95%CI 1.07–1.24; p<0.001) and stroke-metaGRS (HR per SD increment, 1.10; 95%CI 1.02–1.18; p=0.013) were associated with MACE. After full adjustment for conventional vascular risk factors, the association remained significant for CAD-metaGRS (HR=1.10; 95%CI 1.02–1.18; p=0.016) but not for stroke-metaGRS (HR=1.04; 95%CI 0.96–1.12; p=0.338). Neither score was significantly associated with recurrent stroke. For stroke-metaGRS, the HRs were 1.10 (95%CI 0.99–1.22; p=0.072) in the age- and sex-adjusted model and 1.05 (95%CI 0.95–1.17; p=0.344) in the fully adjusted model. Corresponding estimates for CAD-metaGRS were 1.06 (95%CI 0.95–1.17; p=0.324) and 1.02 (95%CI 0.92–1.14; p=0.718), respectively (**Figure 1**).

**Figure 1.**
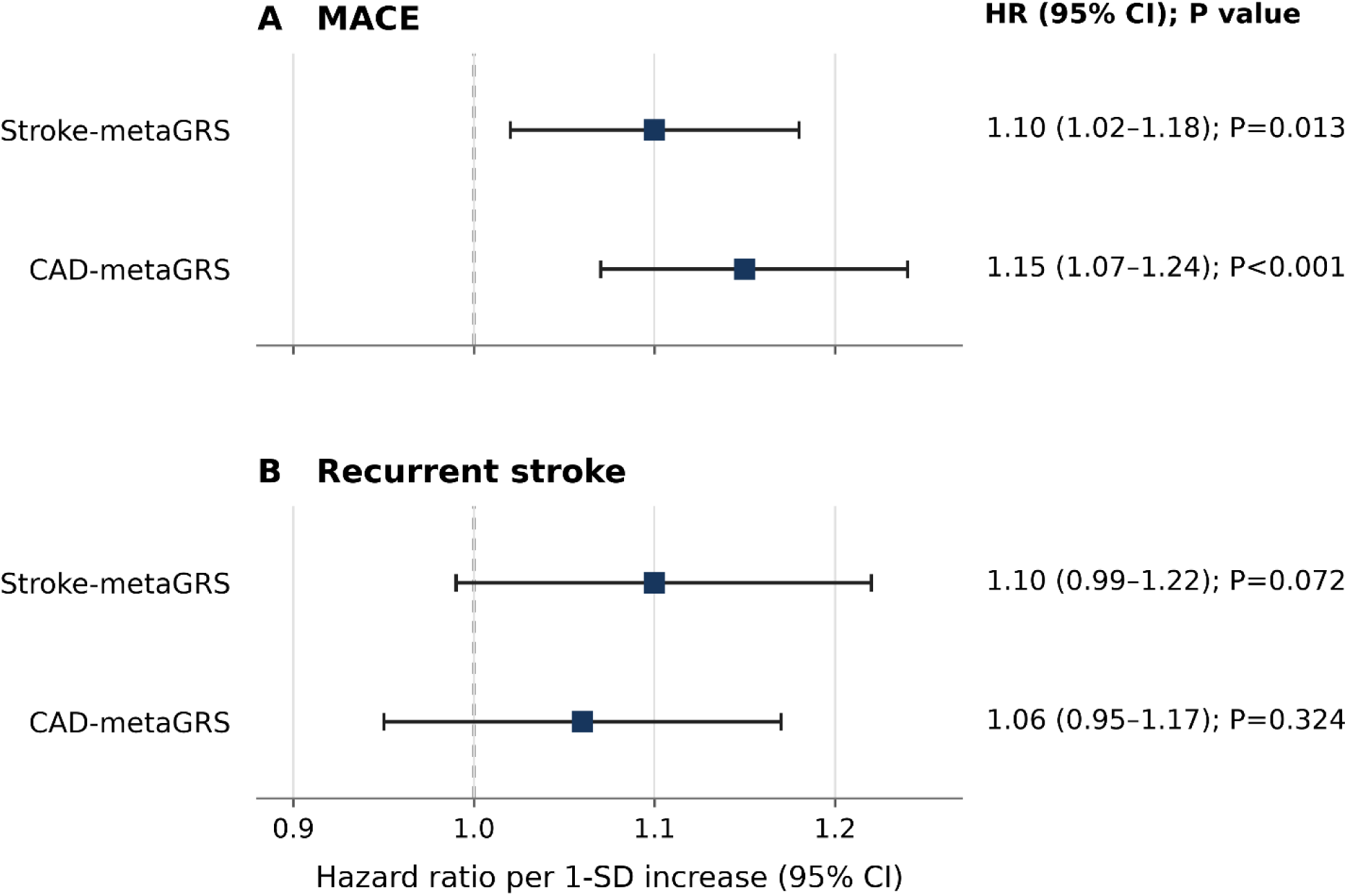
Associations of stroke- and coronary artery disease polygenic risk scores with MACE and recurrent stroke.

Exploratory percentile analyses showed progressively higher MACE risk at more extreme CAD-metaGRS thresholds. Compared with the remaining participants, the age- and sex-adjusted HRs were 1.49 (95%CI 1.20–1.85) for the top 10%, 1.52 (95%CI 1.12–2.05) for the top 5%, and 2.13 (95%CI 1.17–3.87) for the top 1%. After adjustment for vascular risk factors, the corresponding HRs were 1.31 (95%CI 1.05–1.64), 1.37 (95%CI 1.02–1.86), and 1.95 (95%CI 1.07–3.56), respectively.

Similarly, exploratory percentile analyses of the stroke-metaGRS showed progressively larger recurrent-stroke estimates at more extreme thresholds. Compared with the remaining participants, the age- and sex-adjusted HRs were 1.33 (95%CI 0.97–1.84) for the top 10%, 1.77 (95%CI 1.18–2.64) for the top 5%, and 2.69 (95%CI 1.27–5.71) for the top 1%. After adjustment for vascular risk factors, the corresponding HRs were 1.18 (95%CI 0.85–1.63), 1.57 (95%CI 1.04–2.36), and 2.27 (95%CI 1.06–4.86), respectively.

### Associations between polygenic risk scores and vascular risk factor control

**Figure 2** depicts the associations between the stroke-metaGRS and CAD-metaGRS, respectively, and failure to achieve guideline-recommended vascular risk factor targets among stroke survivors. In age- and sex-adjusted analyses, the stroke-metaGRS demonstrated a selective association with glycaemic control. Specifically, higher stroke-metaGRS values were associated with significantly greater odds of failing to achieve HbA1c targets of both <6.5% and <7.0%, with these associations remaining robust after adjustment for age and sex (adjusted OR 1.315, 95%CI 1.167–1.481, p<0.001, and adjusted OR 1.217, 95%CI 1.054–1.406, p=0.007, respectively). In contrast, stroke-metaGRS was not associated with blood-pressure or LDL-C threshold exceedance; for LDL-C >70 mg/dL, the adjusted OR was 0.874 (95%CI 0.754–1.013; p=0.074). The CAD-metaGRS exhibited a complementary pattern of associations. In both univariable and multivariable analyses, higher CAD-metaGRS values were independently associated with increased odds of failing to achieve the most stringent LDL-C target (<70 mg/dL) (adjusted OR 1.22, 95%CI 1.04–1.42, p=0.012), whereas no significant associations were identified for the less stringent LDL-C thresholds (<100 or <130 mg/dL). Moreover, the CAD-metaGRS was consistently associated with poorer glycaemic control, including failure to achieve HbA1c targets of <6.5% (adjusted OR 1.31, 95%CI 1.16–1.48, p<0.001) and <7.0% (adjusted OR 1.185, 95%CI 1.021–1.376, p=0.026). No significant associations were observed between the CAD-metaGRS and systolic or diastolic blood pressure control.

**Figure 2.**
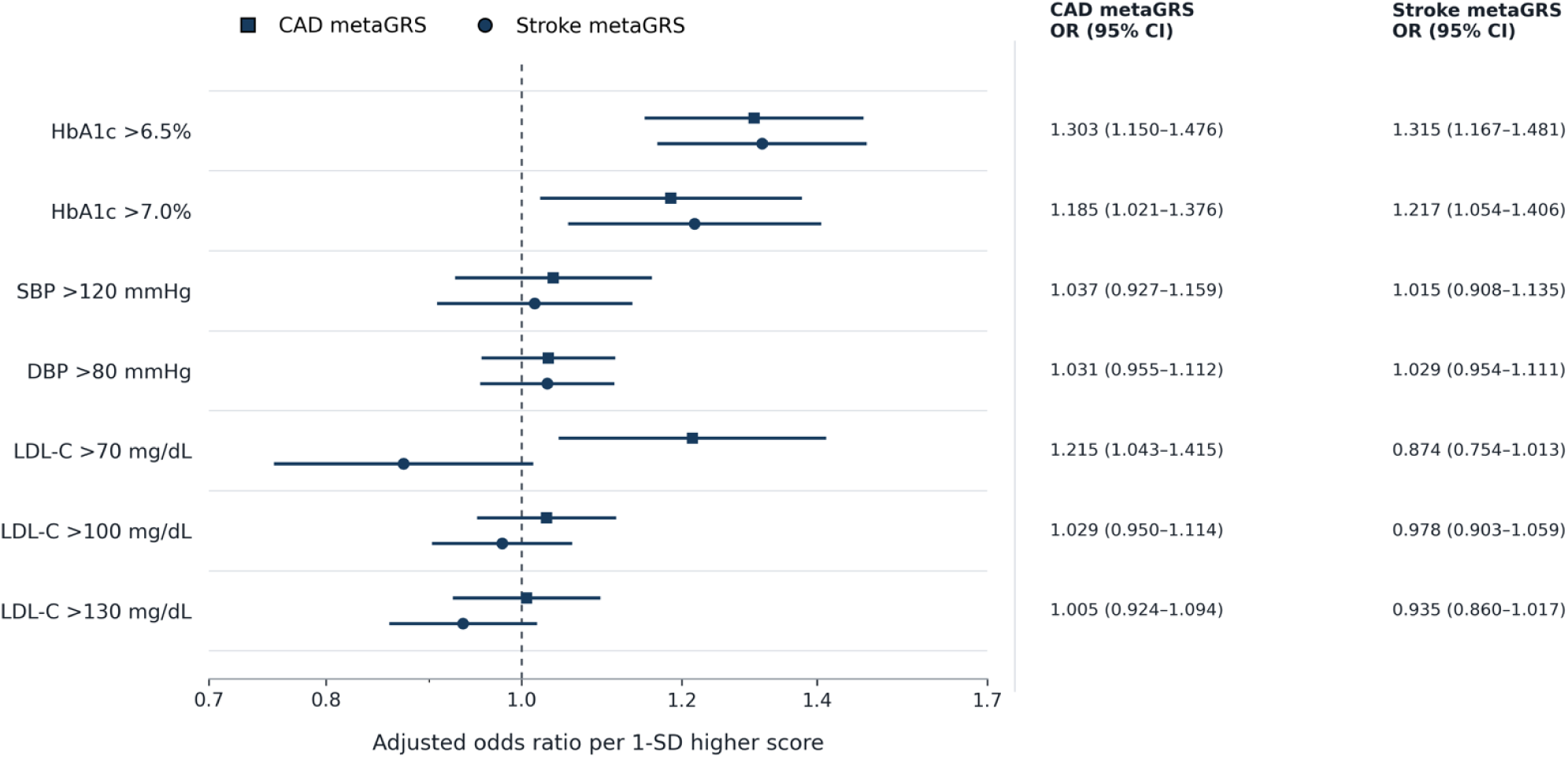
Associations of CAD- and stroke-metaGRSs with baseline cardiometabolic threshold exceedance.

### Mediation of Polygenic Risk by Vascular Risk Factor Control

At the primary 10-year horizon, a 1-SD higher CAD-metaGRS was associated with a total model-based MACE risk difference of 2.33 percentage points (95%CI 0.90–3.83; p=0.002) in the continuous-HbA1c model. The estimated interventional indirect component through continuous HbA1c was 0.39 percentage points (95%CI 0.21–0.59; p<0.001; FDR q=0.004), corresponding to 16.7% (95%CI 8.2–42.1%) of the total association. When HbA1c was modelled as >6.5%, the indirect risk difference was 0.47 percentage points (95%CI 0.20–0.82; p<0.001; FDR q=0.004), corresponding to 19.3% (95%CI 7.9–49.0%) of the total association. Indirect components through HbA1c >7.0% and continuous SBP were nominally supported but did not remain significant after FDR correction (0.22 percentage points; 95%CI 0.02–0.51; p=0.023; q=0.063; and 0.13 percentage points; 95%CI 0.01–0.29; p=0.020; q=0.063, respectively). No evidence of indirect associations through DBP or LDL-C was observed (**Supplemental Table 3**).

For stroke-metaGRS and recurrent stroke, the 10-year total risk difference in the continuous-HbA1c model was 0.82 percentage points (95%CI −0.34 to 2.03; p=0.175), whereas the estimated indirect component through continuous HbA1c was 0.20 percentage points (95%CI 0.10–0.31; p<0.001; FDR q=0.004). The indirect components through HbA1c >6.5% and >7.0% were nominally significant before FDR correction (0.20 percentage points; 95%CI 0.04–0.40; p=0.010; q=0.055; and 0.10 percentage points; 95%CI 0.001–0.25; p=0.044; q=0.161, respectively). No indirect association was supported for the remaining blood-pressure or LDL-C specifications (**Supplemental Table 3**).

The inferential pattern was unchanged at 5 and 15 years; however, the 15-year estimates were considered extrapolative and model-dependent because only 129 participants remained at risk for MACE and 166 for recurrent stroke at that horizon (**Supplemental Table 4**).

### Performance of the Clinical Reference and Integrated metaGRS Models

The clinical reference models demonstrated moderate discrimination for both outcomes, with higher C-indices for MACE than for recurrent stroke. For MACE, the age- and sex-adjusted model achieved a Harrell C-index of 0.622 (SE=0.010), which increased to 0.627 (SE=0.010) after incorporating the CAD-metaGRS. The resulting change in discrimination was small and not statistically significant (ΔC=0.0055; SE=0.0033; 95%CI, −0.0011 to 0.0120; p=0.100), although the LRT indicated improved relative model fit (χ²₁=13.13; p<0.001). The fully adjusted clinical reference model achieved a C-index of 0.677 (SE=0.010), increasing marginally to 0.680 (SE=0.010) after addition of the CAD-metaGRS (ΔC=0.0027; SE=0.0016; 95%CI −0.0003 to 0.0058; p=0.081); nevertheless, the improvement in model fit remained statistically significant (χ²₁=5.74; p=0.017). For recurrent stroke, the age- and sex-adjusted model yielded a C-index of 0.589 (SE=0.015), increasing to 0.595 (SE=0.015) after addition of the stroke-metaGRS (ΔC=0.0051; SE=0.0053; 95%CI −0.0053 to 0.0154; p=0.337), without significant improvement in model fit (χ²₁=3.19; p=0.074). Similarly, the fully adjusted clinical reference model achieved a C-index of 0.630 (SE=0.015), which remained essentially unchanged after incorporating the stroke-metaGRS (C-index=0.631; SE=0.015; ΔC=0.0012; SE=0.0020; 95%CI −0.0028 to 0.0051; p=0.564), with no evidence of improved model fit (χ²₁=0.89; p=0.346). Overall, established clinical and demographic variables accounted for most of the discriminatory performance. The CAD-metaGRS improved relative model fit for MACE but did not significantly improve discrimination, whereas the stroke-metaGRS improved neither model fit nor discrimination for recurrent stroke (**Supplemental Figure 5**).

## Discussion

In this large, well-characterized cohort of stroke survivors drawn from the UK Biobank, we demonstrated that both the polygenic scores for stroke and coronary artery disease independently predicted recurrent vascular events and vascular risk factor control. These findings provide further evidence that polygenic predisposition to atherosclerotic and cardiometabolic disease continues to influence vascular prognosis long after an index vascular event. They also provide potentially actionable insights for more intensive vascular risk factor monitoring among individuals with high polygenic scores.

Our results are consistent with and substantially extend the emerging evidence supporting the role of polygenic risk scores in secondary stroke prevention. Recent work by Han et al.^20^ demonstrated that polygenic risk scores originally developed for incident IS remain associated with recurrent stroke and subsequent coronary heart disease among stroke survivors, indicating that inherited susceptibility continues to influence vascular prognosis despite contemporary secondary prevention. Likewise, Wu et al.^21^ showed that polygenic susceptibility to hyperlipidemia was associated with poorer LDL cholesterol control and higher risks of recurrent stroke and coronary events, supporting the concept that inherited cardiometabolic susceptibility contributes not only to recurrent vascular outcomes but also to failure to achieve optimal risk factor control. More recently, Kojima et al.^22^ developed and validated a stroke-specific metaGRS that independently predicted recurrent stroke in a Japanese cohort, demonstrating that genetic risk remains clinically relevant across ancestries and healthcare systems and supporting the robustness and generalizability of polygenic risk stratification in secondary prevention. Our study extends these observations in several important respects. First, we simultaneously evaluated complementary stroke- and CAD-specific metaGRSs, demonstrating distinct but clinically relevant associations with recurrent stroke and MACE, respectively. Second, beyond outcome prediction, we examined the relationship between polygenic burden and the achievement of guideline-recommended glycemic, lipid, and blood pressure targets. Finally, by incorporating formal mediation analyses, we provide mechanistic evidence that inherited vascular susceptibility is at least partly expressed through modifiable cardiometabolic pathways, particularly glycemic regulation, thereby extending polygenic risk assessment from prognostic stratification toward biological characterization and precision secondary prevention.

A key observation was the graded, dose-dependent increase in event risk across ascending percentiles of the CAD-metaGRS, culminating in an age- and sex-adjusted hazard ratio of 2.13 and a fully adjusted hazard ratio of 1.95 among individuals in the highest one percent of the genetic risk distribution. This monotonic gradient delineates a small but clinically meaningful subgroup of stroke survivors whose inherited risk is markedly amplified. The persistence of excess risk at these extreme percentiles, despite contemporary pharmacologic therapy and routine clinical follow-up, suggests that polygenic burden captures a dimension of vascular vulnerability that is not fully reflected by conventional clinical risk factors. Recognition of this genetically high-risk subgroup may therefore justify more personalized surveillance, stricter treatment targets, and earlier therapeutic escalation. However, this subgroup included only 28 participants, and the confidence intervals were wide. These findings should therefore be considered hypothesis-generating and require validation before extreme polygenic-risk thresholds can guide surveillance or treatment intensification. A similar pattern was observed for stroke-metaGRS and recurrent stroke.

Our mediation analyses further provide mechanistic insight into how inherited susceptibility translates into recurrent vascular events. Glycemic control consistently emerged as the predominant mediator, whereas LDL cholesterol and blood pressure contributed comparatively little, indicating that genetic liability may be preferentially expressed through dysglycemic pathways. This observation is biologically plausible and aligns with accumulating genomic evidence linking cardiometabolic polygenic susceptibility to insulin resistance, vascular aging, endothelial dysfunction, chronic inflammation, and accelerated atherothrombosis.^23,24^ The present findings also complement those of Wu et al.,^21^ who demonstrated that genetic susceptibility to hyperlipidemia is associated with poorer lipid control among stroke survivors. Whereas Wu et al.^21^ focused on a lipid-specific polygenic profile, our analyses simultaneously evaluated cerebrovascular and coronary polygenic susceptibility while formally quantifying the relative contributions of glycemic, blood pressure, and lipid pathways through mediation analysis. The observation that glycemic regulation consistently emerged as the predominant mediator, whereas LDL cholesterol and blood pressure accounted for comparatively little of the observed genetic effect, suggests that inherited vascular susceptibility may preferentially operate through metabolic rather than isolated lipid or hemodynamic pathways. Moreover, the persistence of significant direct effects for the CAD-metaGRS indicates that additional mechanisms, including inflammation, thrombosis, plaque instability, or vascular remodeling, may contribute to recurrent cardiovascular events beyond conventional cardiometabolic pathways. Conversely, the largely indirect effects observed for the stroke-metaGRS support the hypothesis that glycemic dysregulation represents an important intermediate phenotype linking inherited cerebrovascular susceptibility to stroke recurrence.

Taken together, these findings suggest that the adverse metabolic phenotype observed among genetically high-risk stroke survivors is not merely a consequence of behavioral factors or treatment variability but may instead reflect an intrinsic, genetically determined predisposition to impaired cardiometabolic homeostasis. Polygenic risk therefore functions not only as a prognostic marker but also as a surrogate for underlying biological vulnerability, providing a rationale for more individualized preventive strategies.^25^

From a clinical perspective, our findings have several important implications. First, polygenic risk scores can identify stroke survivors whose residual vascular risk remains disproportionately high despite otherwise favorable clinical profiles and contemporary secondary prevention. Second, the mechanistic pathways identified, particularly the prominent role of glycemic regulation, highlight potentially modifiable therapeutic targets. Stroke survivors with high polygenic burden may benefit from closer glycemic surveillance, earlier implementation of intensive metabolic management, and consideration of therapies with established cardiovascular benefit, although specific pharmacological recommendations require prospective validation.

## Limitations and Future Directions

Several limitations warrant consideration. First, the UK Biobank cohort, though large and deeply phenotyped, is not population-representative and predominantly of European ancestry, potentially limiting generalizability to other ancestries. Second, medication adherence, treatment intensity, and post-stroke rehabilitation adherence, factors that could influence recurrent risk, were not fully captured. Third, the observed increases in discrimination remain small in absolute terms; thus, genetic risk scores should complement rather than replace established risk markers. Finally, our mediation analyses, though suggestive of causal pathways, are observational and can only be used to guide future research focused on interventional designs.

Future studies should address these shortcomings, validating these findings in multi-ethnic stroke cohorts and evaluating whether integrating polygenic risk into dynamic risk prediction models improves clinical outcomes. Trials exploring genotype-informed intensification of metabolic and vascular management in genetically high-risk survivors could establish causal utility. Beyond prediction, functional genomic analyses may elucidate the biological pathways mediating polygenic risk, opening new therapeutic avenues targeting metabolic–vascular cross-talk.

## Conclusions

In a cohort of stroke survivors, polygenic risk scores for coronary artery disease and stroke were associated with residual vascular risk and vascular risk factor control. In particular, a polygenic score for coronary artery disease was associated with glycemic control, which appeared to mediate its association with future cardiovascular events. These findings warrant further research but suggest that polygenic risk scores may help identify patients who could benefit from more intensive cardiometabolic risk factor management as part of secondary stroke prevention.

## Data Availability

All data produced in the present work are contained in the manuscript

---

**Supplemental Table 1.** Phenotype definitions, source fields, and diagnostic codes.

| Phenotype | Source | Field/coding system | Definition or codes | Timing |
| --- | --- | --- | --- | --- |
| Prevalent stroke | Self-report | UK Biobank field 20002 | 1081 (stroke), 1583 (ischemic stroke), 1491 (brain hemorrhage) | Before baseline |
|  | Hospital/death records | ICD-10; ICD-9 | I60–I64; 430–434 and 436 | Before baseline |
| Nonfatal myocardial infarction component of MACE | Hospital inpatient records | ICD-10 | I21–I22 | After baseline; first qualifying event |
| Stroke component of MACE and recurrent stroke | Hospital/death records | ICD-10 | I60–I64 | After baseline; first qualifying event |
| Cardiovascular death component of MACE | Death registry | Underlying-cause ICD-10 | I00–I99 | After baseline |
| LDL-C off target | Baseline biochemistry | UK Biobank field 30780 | >70, >100, or >130 mg/dL | Baseline |
| Glycated hemoglobin off target | Baseline biochemistry | UK Biobank field 30750 | >6.5% or >7.0% | Baseline |
| Blood pressure off target | Baseline physical measures | UK Biobank fields 4080/93 (SBP) and 4079/94 (DBP) | SBP >120 mm Hg or DBP >80 mm Hg | Baseline |

**Supplemental Table 2.** Baseline clinical, anthropometric, and biochemical characteristics of stroke survivors from the UK Biobank according to MACE and recurrent stroke status.

| Variable | No recurrent stroke<br>N = 2,350 (87.0%) | Recurrent stroke<br>N = 351 (13.0%) | p-value | No MACE<br>N = 1,970 (72.9%) | MACE<br>N = 731 (27.1%) | p-value |
| --- | --- | --- | --- | --- | --- | --- |
| Sex |  |  | 0.5 |  |  | <0.001 |
| Female, n (%) | 906 (39%) | 129 (37%) |  | 803 (41%) | 232 (32%) |  |
| Male, n (%) | 1,444 (61%) | 222 (63%) |  | 1,167 (59%) | 499 (68%) |  |
| Age (years) | 59.5 (7.2) | 61.9 (6.4) | <0.001 | 58.9 (7.2) | 62.3 (6.1) | <0.001 |
| Waist circumference (cm) | 95.6 (13.6) | 97.9 (13.3) | 0.001 | 94.9 (13.5) | 98.7 (13.6) | <0.001 |
| Hip circumference (cm) | 104.5 (9.5) | 105.7 (9.5) | 0.032 | 104.5 (9.3) | 105.3 (10.1) | 0.2 |
| BMI (kg/m<sup>2</sup>) | 28.6 (5.0) | 29.2 (5.0) | 0.013 | 28.5 (5.0) | 29.2 (5.2) | 0.001 |
| Systolic blood pressure (mmHg) | 141.2 (19.7) | 144.0 (21.4) | 0.011 | 140.9 (19.2) | 143.4 (22.0) | 0.007 |
| Diastolic blood pressure (mmHg) | 82.2 (11.2) | 81.9 (11.9) | 0.574 | 82.5 (10.9) | 81.2 (12.2) | 0.007 |
| LDL-C (mmol/L) | 3.0 (0.9) | 2.8 (1.0) | <0.001 | 3.0 (0.9) | 2.8 (0.9) | <0.001 |
| (mg/dL) | 115.3 (34.1) | 109.7 (36.8) |  | 116.5 (34.6) | 109.4 (33.9) |  |
| HDL-C (mmol/L) | 1.3 (0.4) | 1.3 (0.4) | 0.020 | 1.4 (0.4) | 1.3 (0.4) | <0.001 |
| (mg/dL) | 51.8 (14.1) | 50.2 (14.0) |  | 52.3 (14.0) | 49.5 (14.2) |  |
| Triglycerides (mmol/L) | 1.8 (1.1) | 1.9 (1.0) | 0.435 | 1.8 (1.1) | 1.9 (1.0) | 0.073 |
| (mg/dL) | 162.6 (95.3) | 163.9 (87.2) |  | 161.5 (94.9) | 166.3 (92.7) |  |
| Total cholesterol (mmol/L) | 4.9 (1.2) | 4.7 (1.3) | <0.001 | 5.0 (1.2) | 4.7 (1.2) | <0.001 |
| (mg/dL) | 190.8 (45.7) | 182.7 (49.3) |  | 192.6 (46.0) | 182.0 (46.1) |  |
| eGFRcr-cys (mL/min/1.73 m<sup>2</sup>) | 87.4 (17.5) | 83.6 (18.2) | <0.001 | 89.2 (16.3) | 80.7 (19.5) | <0.001 |
| Cystatin C (mg/L) | 1.0 (0.3) | 1.0 (0.3) | <0.001 | 1.0 (0.2) | 1.1 (0.3) | <0.001 |
| Creatinine (μmol/L) | 78.5 (24.9) | 80.5 (27.4) | 0.5 | 76.9 (20.8) | 83.9 (34.1) | <0.001 |
| (mg/dL) | 0.9 (0.3) | 0.9 (0.3) |  | 0.9 (0.2) | 0.9 (0.4) |  |
| HbA1c (mmol/mol) | 38.6 (13.1) | 42.0 (12.5) | <0.001 | 38.0 (13.1) | 42.0 (12.5) | <0.001 |
| (%) | 5.7 (1.2) | 6.0 (1.1) |  | 5.6 (1.2) | 6.0 (1.1) |  |
| Former smoking, n (%) | 1,050 (45%) | 159 (45%) | 0.8 | 862 (44%) | 347 (47%) | 0.085 |
| Current smoking, n (%) | 355 (15%) | 54 (15%) | 0.9 | 270 (14%) | 139 (19%) | <0.001 |
| Antihypertensive medication, n (%) | 1,387 (59%) | 234 (67%) | 0.006 | 1,118 (57%) | 503 (69%) | <0.001 |
| Statin use, n (%) | 1,468 (62%) | 251 (72%) | 0.001 | 1,177 (60%) | 542 (74%) | <0.001 |
| Diabetes, n (%) | 278 (12%) | 82 (23%) | <0.001 | 189 (9.6%) | 171 (23%) | <0.001 |
| Hypertension, n (%) | 1,536 (65%) | 255 (73%) | 0.007 | 1,234 (63%) | 557 (76%) | <0.001 |

**Supplemental Table 3.** Ten-year interventional mediation of genetic risk-score associations with MACE and recurrent stroke through metabolic and vascular mediators. RD: risk difference.

| Mediator | Total effect, RD pp (95% CI) | Direct effect, RD pp (95% CI) | Indirect effect, RD pp (95% CI) | p-value |
| --- | --- | --- | --- | --- |
| **CAD-metaGRS to MACE** |  |  |  |  |
| HbA1c (continuous, per SD) | 2.329 (0.902, 3.832) | 1.941 (0.528, 3.393) | 0.388 (0.207, 0.590) | <0.001 |
| HbA1c >6.5% | 2.419 (0.941, 3.953) | 1.951 (0.495, 3.467) | 0.468 (0.200, 0.819) | <0.001 |
| HbA1c >7.0% | 2.438 (0.960, 3.983) | 2.222 (0.763, 3.712) | 0.216 (0.022, 0.505) | 0.023 |
| SBP (continuous, per SD) | 2.601 (1.153, 4.091) | 2.476 (1.041, 3.952) | 0.125 (0.012, 0.295) | 0.020 |
| SBP >120 mmHg | 2.569 (1.107, 4.079) | 2.562 (1.092, 4.079) | 0.008 (-0.033, 0.068) | 0.769 |
| DBP (continuous, per SD) | 2.544 (1.096, 4.072) | 2.532 (1.075, 4.054) | 0.011 (-0.044, 0.100) | 0.681 |
| DBP >80 mmHg | 2.471 (1.026, 3.992) | 2.480 (1.032, 3.992) | -0.009 (-0.070, 0.042) | 0.784 |
| LDL-C (continuous, per SD) | 2.589 (1.135, 4.097) | 2.594 (1.136, 4.098) | -0.005 (-0.072, 0.058) | 0.886 |
| LDL-C >70 mg/dL | 2.517 (1.071, 4.049) | 2.519 (1.067, 4.038) | -0.003 (-0.100, 0.101) | 0.970 |
| LDL-C >100 mg/dL | 2.517 (1.092, 4.029) | 2.534 (1.104, 4.048) | -0.017 (-0.096, 0.041) | 0.617 |
| LDL-C >130 mg/dL | 2.523 (1.076, 4.032) | 2.524 (1.075, 4.031) | -0.001 (-0.042, 0.043) | 0.999 |
| **stroke-metaGRS to recurrent stroke** |  |  |  |  |
| HbA1c (continuous, per SD) | 0.819 (-0.344, 2.034) | 0.622 (-0.510, 1.801) | 0.197 (0.096, 0.308) | <0.001 |
| HbA1c >6.5% | 0.862 (-0.324, 2.120) | 0.665 (-0.529, 1.918) | 0.196 (0.043, 0.397) | 0.010 |
| HbA1c >7.0% | 0.877 (-0.293, 2.128) | 0.779 (-0.403, 2.023) | 0.098 (0.001, 0.255) | 0.044 |
| SBP (continuous, per SD) | 0.937 (-0.217, 2.148) | 0.859 (-0.277, 2.073) | 0.078 (-0.009, 0.205) | 0.086 |
| SBP >120 mmHg | 0.946 (-0.226, 2.197) | 0.947 (-0.221, 2.197) | -0.002 (-0.039, 0.028) | 0.954 |
| DBP (continuous, per SD) | 0.941 (-0.227, 2.164) | 0.927 (-0.235, 2.152) | 0.014 (-0.052, 0.100) | 0.700 |
| DBP >80 mmHg | 0.929 (-0.242, 2.163) | 0.932 (-0.242, 2.172) | -0.003 (-0.046, 0.036) | 0.937 |
| LDL-C (continuous, per SD) | 0.973 (-0.193, 2.230) | 0.961 (-0.234, 2.218) | 0.011 (-0.059, 0.117) | 0.747 |
| LDL-C >70 mg/dL | 0.951 (-0.229, 2.199) | 0.951 (-0.224, 2.194) | 0.000 (-0.067, 0.064) | 0.981 |
| LDL-C >100 mg/dL | 0.997 (-0.159, 2.211) | 0.987 (-0.166, 2.199) | 0.010 (-0.034, 0.073) | 0.687 |
| LDL-C >130 mg/dL | 0.980 (-0.202, 2.220) | 0.992 (-0.206, 2.243) | -0.012 (-0.073, 0.044) | 0.645 |

**Supplemental Table 4.** Indirect risk differences across the 5-, 10-, and 15-year horizons.

| Mediator | 5-year indirect RD, pp (95% CI) | 10-year indirect RD, pp (95% CI) | 15-year indirect RD, pp (95% CI)* |
| --- | --- | --- | --- |
| **CAD-metaGRS to MACE** |  |  |  |
| HbA1c (continuous, per SD) | 0.209 (0.110, 0.321) | 0.388 (0.207, 0.590) | 0.517 (0.278, 0.783) |
| HbA1c >6.5% | 0.267 (0.112, 0.476) | 0.468 (0.200, 0.819) | 0.586 (0.254, 1.010) |
| HbA1c >7.0% | 0.125 (0.012, 0.300) | 0.216 (0.022, 0.505) | 0.268 (0.028, 0.612) |
| SBP (continuous, per SD) | 0.068 (0.006, 0.161) | 0.125 (0.012, 0.295) | 0.167 (0.016, 0.390) |
| SBP >120 mmHg | 0.004 (-0.017, 0.036) | 0.008 (-0.033, 0.068) | 0.011 (-0.044, 0.093) |
| DBP (continuous, per SD) | 0.006 (-0.024, 0.055) | 0.011 (-0.044, 0.100) | 0.015 (-0.059, 0.136) |
| DBP >80 mmHg | -0.005 (-0.038, 0.023) | -0.009 (-0.070, 0.042) | -0.012 (-0.092, 0.056) |
| LDL-C (continuous, per SD) | -0.003 (-0.038, 0.031) | -0.005 (-0.072, 0.058) | -0.007 (-0.095, 0.078) |
| LDL-C >70 mg/dL | -0.001 (-0.057, 0.054) | -0.003 (-0.100, 0.101) | -0.004 (-0.128, 0.135) |
| LDL-C >100 mg/dL | -0.009 (-0.052, 0.022) | -0.017 (-0.096, 0.041) | -0.022 (-0.127, 0.055) |
| LDL-C >130 mg/dL | 0.000 (-0.022, 0.023) | -0.001 (-0.042, 0.043) | -0.001 (-0.056, 0.058) |
| **stroke-metaGRS to recurrent stroke** |  |  |  |
| HbA1c (continuous, per SD) | 0.109 (0.053, 0.173) | 0.197 (0.096, 0.308) | 0.271 (0.132, 0.424) |
| HbA1c >6.5% | 0.111 (0.023, 0.227) | 0.196 (0.043, 0.397) | 0.266 (0.058, 0.531) |
| HbA1c >7.0% | 0.056 (0.001, 0.146) | 0.098 (0.001, 0.255) | 0.133 (0.002, 0.342) |
| SBP (continuous, per SD) | 0.044 (-0.005, 0.116) | 0.078 (-0.009, 0.205) | 0.108 (-0.012, 0.281) |
| SBP >120 mmHg | -0.001 (-0.022, 0.016) | -0.002 (-0.039, 0.028) | -0.002 (-0.051, 0.039) |
| DBP (continuous, per SD) | 0.008 (-0.029, 0.056) | 0.014 (-0.052, 0.100) | 0.019 (-0.072, 0.139) |
| DBP >80 mmHg | -0.001 (-0.025, 0.020) | -0.003 (-0.046, 0.036) | -0.004 (-0.063, 0.050) |
| LDL-C (continuous, per SD) | 0.006 (-0.032, 0.066) | 0.011 (-0.059, 0.117) | 0.016 (-0.080, 0.162) |
| LDL-C >70 mg/dL | 0.000 (-0.037, 0.036) | 0.000 (-0.067, 0.064) | 0.000 (-0.094, 0.088) |
| LDL-C >100 mg/dL | 0.005 (-0.019, 0.042) | 0.010 (-0.034, 0.073) | 0.013 (-0.047, 0.100) |
| LDL-C >130 mg/dL | -0.007 (-0.041, 0.025) | -0.012 (-0.073, 0.044) | -0.017 (-0.101, 0.061) |

\*The 15-year estimates are extrapolative because fewer than 7% of participants remained under observation beyond 15 years.

**Supplemental Figure 1.**
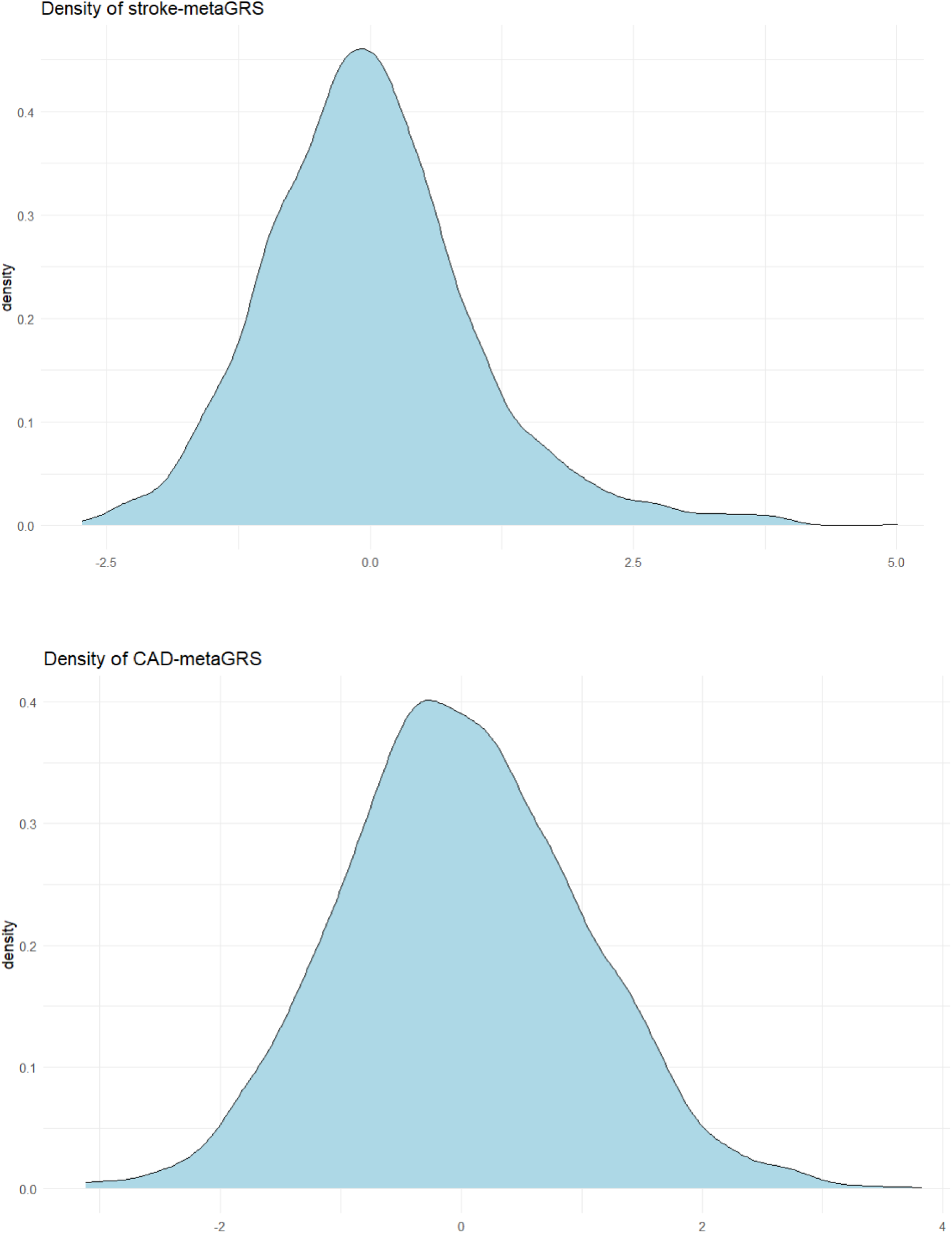
Distribution of the stroke- and CAD-metaGRS in the study population.

**Supplemental Figure 2.**
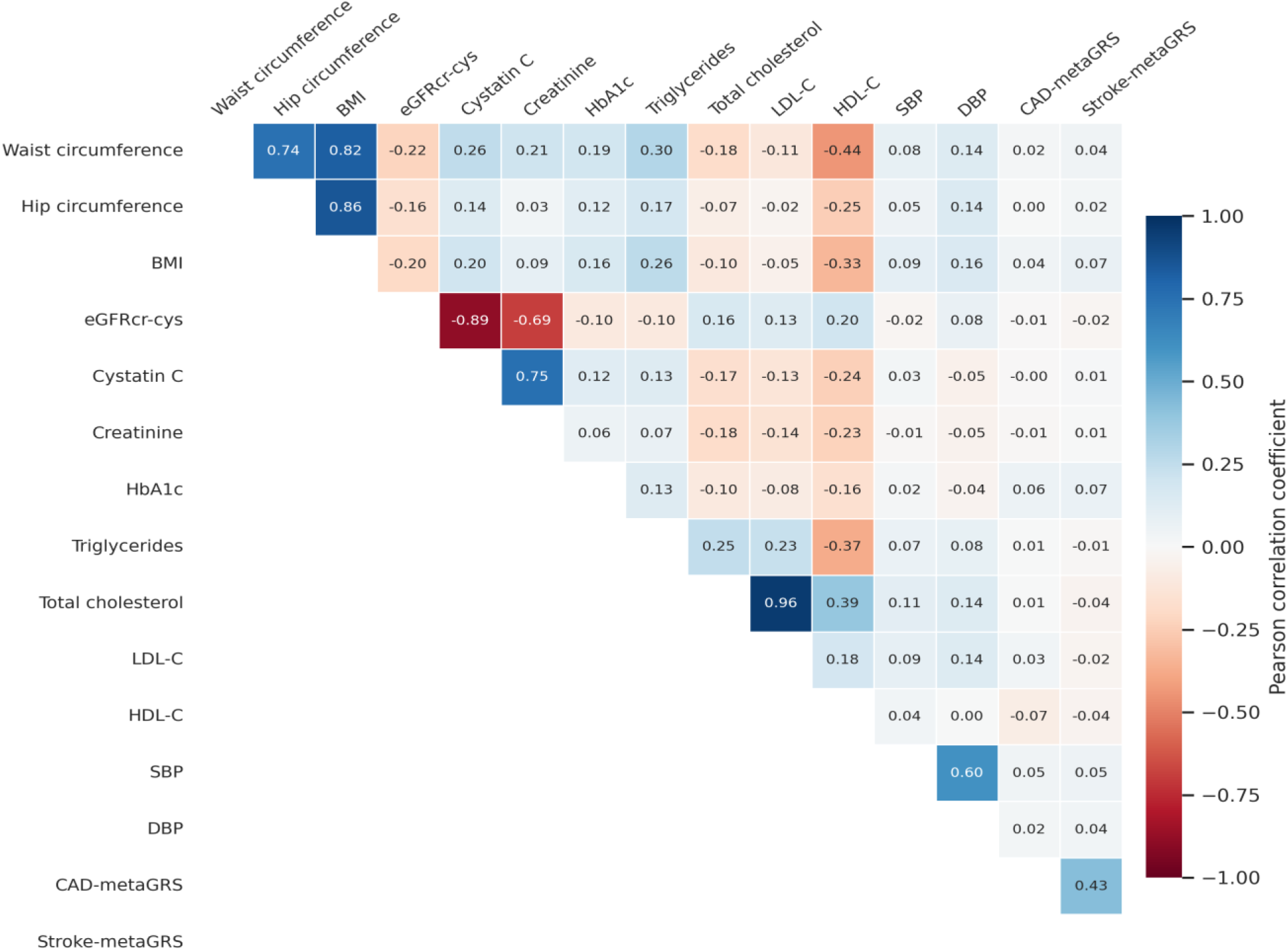
Correlation heatmap of anthropometric, biochemical, and polygenic variables in stroke survivors. Cells show Pearson correlation coefficients; blue denotes positive and red negative correlations.

**Supplemental Figure 3.**
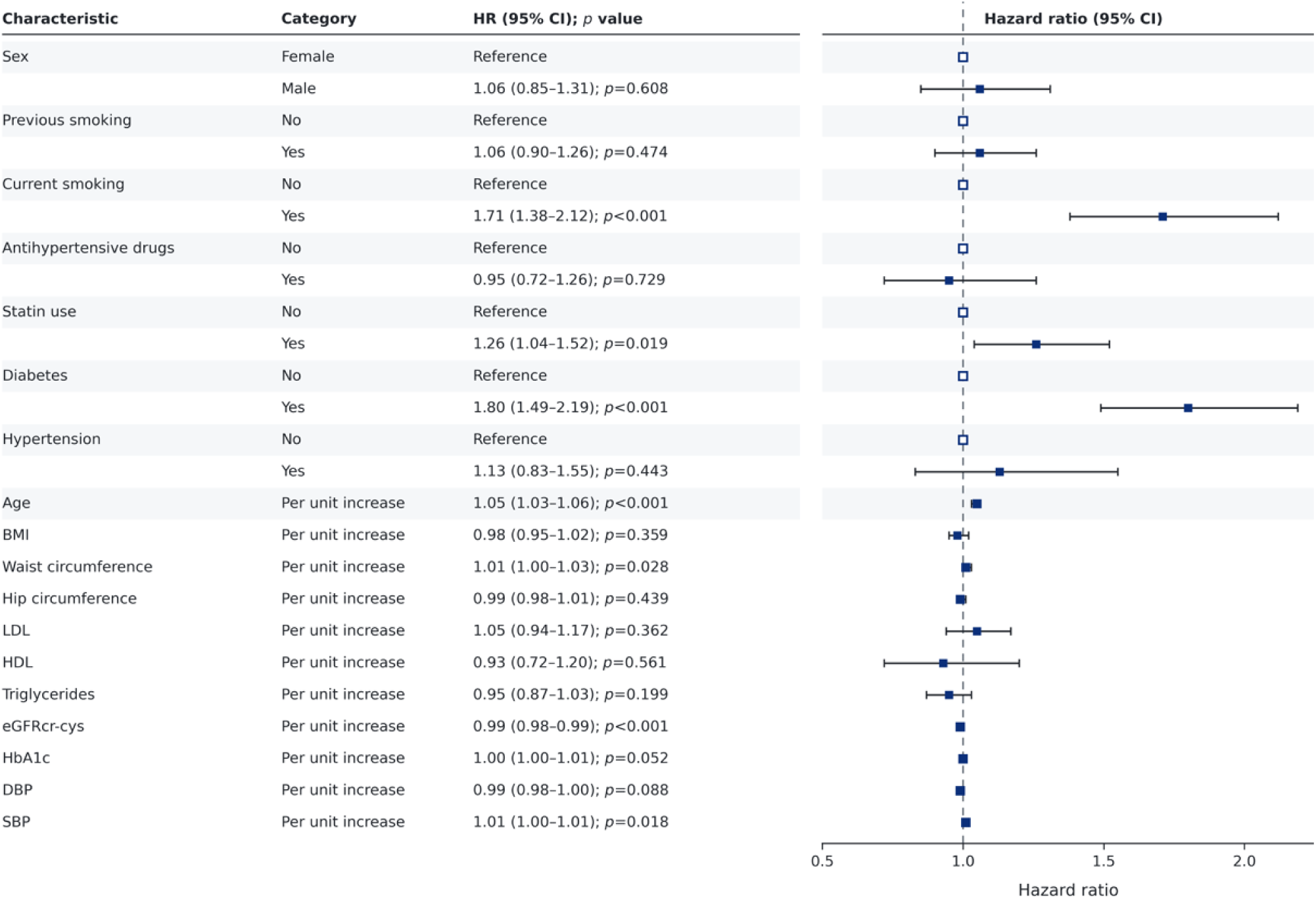
Multivariable Cox proportional hazards analysis for future MACE events in stroke survivors.

**Supplemental Figure 4.**
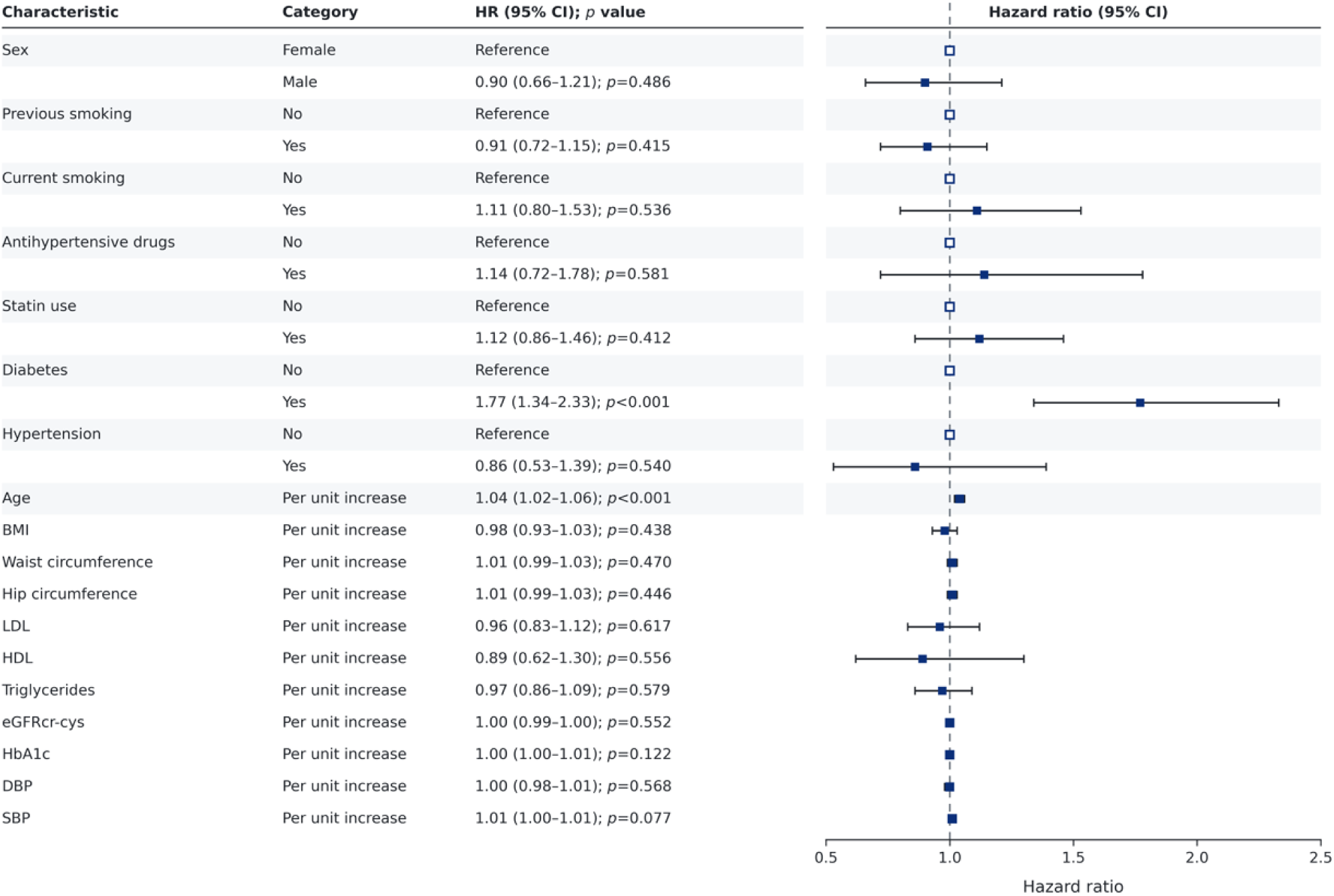
Multivariable Cox proportional hazards analysis for recurrent stroke.

**Supplemental Figure 5.**
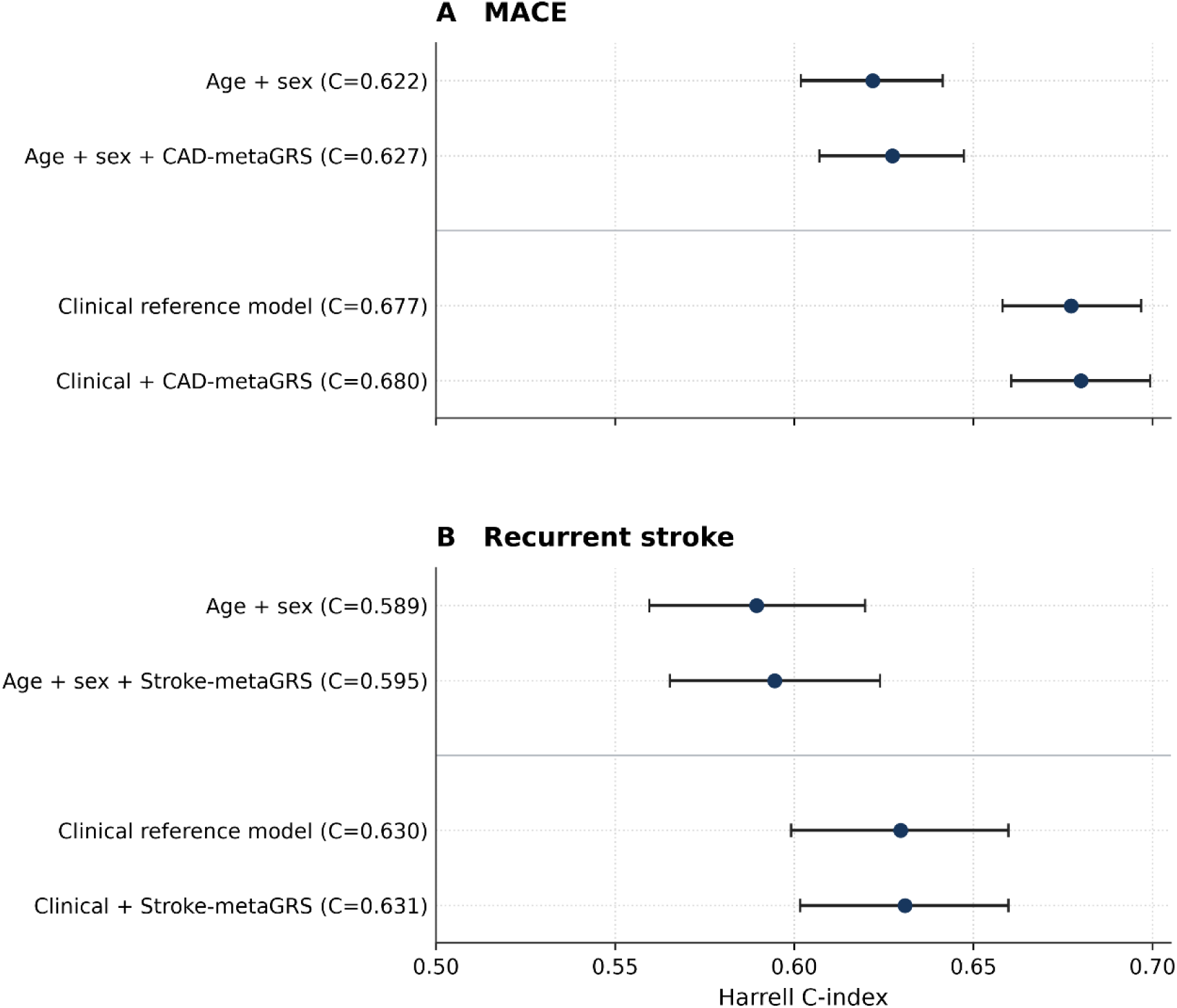
Model discrimination for MACE and recurrent stroke after incorporating polygenic risk.

